# The p–fr–nb triplet: a unified framework for statistical fragility and robustness across clinical study designs

**DOI:** 10.1101/2025.11.20.25340684

**Authors:** Thomas F. Heston

## Abstract

Clinical studies commonly report p-values but rarely quantify how stable those p-values are or how far the observed data lie from the point representing no effect. This study introduces a unified framework that evaluates statistical significance, fragility, and neutrality distance across three standard clinical data structures: single-arm binomial outcomes, two-arm binary outcomes, and continuous two-group outcomes. The objective was to determine whether reporting these three components together can improve the interpretation of clinical research results. Using previously published summary statistics, we calculated significance, fragility, and neutrality distance for representative examples from each design category. The framework applies the diagnostic fragility quotient and a proportion-based neutrality measure for single-arm benchmarks; the global fragility quotient and risk quotient for two-arm binary outcomes; and the continuous fragility scale and meaningful change index for mean comparisons. Across all examples, the triplet revealed patterns that were not detectable with p-values or effect sizes alone. Some statistically significant findings were highly fragile or close to neutrality despite appearing reliable. At the same time, some non-significant results showed meaningful separation from the no-effect state despite stable p-values. These findings highlight how statistical significance, decision stability, and distance from neutrality represent distinct dimensions of evidence that can diverge in clinically important ways. This triplet provides a concise, generalizable summary of evidence quality that enhances transparency and reduces misinterpretation across a broad range of study designs.

## Introduction

Statistical inference in clinical research relies heavily on p-values, confidence intervals, and effect sizes [1]. Although these measures characterize statistical significance and estimate the magnitude of an effect, none directly quantify the stability of those findings or the geometric position of the observed effect relative to the neutrality boundary that defines “no effect” [2]. The traditional landscape is dominated by significance testing, and interpretation frequently collapses into a binary distinction between significant/not significant. Decades of methodological critique—ranging from false-positive findings to concerns about replicability—demonstrate that p-values alone are insufficient for evaluating evidentiary strength or reliability [3–6].

Recent work on fragility has expanded the field by introducing quantitative measures of how easily statistical conclusions can be overturned. Fragility indices and related approaches examine the minimal perturbation needed to reverse statistical significance. Despite their value, applications remain inconsistent, often limited to 2×2 trials, and rarely generalized to continuous or single-arm endpoints [7–11]. Moreover, fragility metrics describe the *stability* of the significance decision, not the *location* of the observed effect relative to its neutrality boundary [12]. As a result, two studies may exhibit similar effect sizes and p-values yet differ markedly in evidentiary strength, depending on how close their results lie to the no-effect state.

A central gap, therefore, is the absence of an integrated perspective that simultaneously captures (1) statistical significance, (2) fragility—movement along the stability axis from fragile to stable, and (3) robustness—movement along the neutrality axis from near-neutrality to far-from-neutrality. These represent distinct dimensions of evidence. Fragility indicates how easily the statistical decision can be reversed. Robustness quantifies the geometric distance between the observed effect and the neutral effect. Without both perspectives, interpretation remains incomplete.

To our knowledge, this is the first study to present a unified evidence-assessment framework that integrates significance, fragility, and neutrality distance across three major clinical endpoint types—single-arm binomial, two-arm binary, and continuous two-group designs—using only published summary statistics. This framework enables a generalizable, multidimensional evaluation of evidentiary strength. The present study demonstrates the use of this triplet across representative examples, computing the p-value, the appropriate fragility metric, and the corresponding neutrality metric for each. We show how these components jointly clarify evidence quality in ways not attainable through traditional statistical reporting alone.

## Materials and methods

The Fragility–Robustness framework assesses evidence quality through three orthogonal dimensions: statistical significance (p), fragility (fr), and robustness (nb).

### Statistical significance (p)

Statistical significance is determined by the p-value, which indicates the likelihood of the observed result under the null hypothesis. Statistical significance is calculated by any generalized method; however, when possible, exact tests are preferred. For example, single-arm outcome studies typically use a one-sided exact binomial test to compare against a benchmark. For two-arm binary outcomes, a two-sided Fisher’s exact test is used. For continuous two-group outcomes, the Welch t-test, which allows unequal variances, is used. The summary statistic is the p-value, which ranges from 0 (an unlikely result under the null hypothesis) to 1 (a highly likely result under the null hypothesis).

### Statistical fragility (fr)

Fragility measures the minimum number of discrete outcome changes needed to cross the p = 0.05 boundary, expressed as a proportion of the relevant sample size. The summary statistic for fragility is “fr” and ranges from 0 (fragile) to 1 (stable).

- Single-arm studies utilize the diagnostic fragility quotient (DFQ) = number of responders that must become non-responders to make p flip significance, divided by total n. It is calculated from the diagnostic fragility index (DFI), which is the number of outcome toggles required to flip statistical significance.
- Two-arm binary outcome studies utilize the global fragility quotient (GFQ) = minimal path-independent cell toggles required to make p > 0.05, divided by total N. The GFQ is calculated from the global fragility index (GFI), which is a label-independent method to determine the minimum number of outcome changes required to flip significance [13].
- Continuous outcomes utilize the continuous fragility quotient (CFQ) = distance of the observed t-statistic from the critical t-value (df-based, α = 0.05 two-sided), normalized to 0–1.

Additional calculation details and other tests to quantify statistical fragility are detailed in the fragility-robustness framework reference document [14].

### Statistical robustness (nb)

Robustness quantifies how far the observed effect lies from neutrality, i.e., the absence of a relationship between the two variables being evaluated. Unlike statistical significance and fragility, it is not based upon probability but rather geometric distance. Based on the Neutrality Boundary Framework (NBF), the summary statistic is “nb” and ranges from 0 (close to neutrality) to 1 (distant from neutrality) [2].

The general equation for NBF statistics is: nb = |T − T_0| / (|T − T_0| + S), where T is the observed estimate, T_0 is neutrality (e.g., benchmark proportion or equality), and S is a sampling variability scale.

Commonly utilized NBF statistics are:

- Single-arm benchmark: Proportion-NBF; scale factor is the standard error of the benchmark proportion.
- Two-arm binary: risk quotient (RQ); distance based on absolute deviation from expected cell counts under independence.
- Continuous two-group: meaningful change index (MeCI); distance of the two means from their weighted midpoint, scaled by pooled standard deviation.

### Interpretation

The interpretation of the p–fr–nb triplet is claim-dependent:

- When claiming an effect exists (p ≤ 0.05): prefer low p, high fr (p-value is stable), high nb (result is distant from neutrality).
- When claiming no effect (p > 0.05): prefer high p, high fr (p-value is stable), low nb (result is close to neutrality).

Similar to the p-value, a wide variety of tests can determine the summary statistic for fragility (fr) and robustness (nb) [14]. Test selection depends upon the underlying data.

The worked examples below used only published summary statistics. No raw patient-level data were collected. Ethics board review is not required for the use of previously published summary statistics. Computations used the Fragility Metrics Toolkit v4.1.0 (Python hosted on Google Colab), which is available as open-source [15].

## Results

### Example 1 — Concordant strong evidence: single-arm benchmark design

A phase 2 trial evaluated olutasidenib in 147 efficacy-evaluable patients with relapsed/refractory mutant IDH1 acute myeloid leukemia. The primary endpoint was complete remission or complete remission with partial hematologic recovery (CR/CRh); the prespecified historical benchmark was 15% [16].

#### Significance

Observed proportion: p̂ = 51/147 = 0.3469. The one-sided exact binomial test (H_0_: p ≤ 0.15): p = 2.65 × 10^−9^ . The observed results are statistically significant, supporting the alternative hypothesis that an effect exists.

#### Fragility (DFQ)

The baseline p-value is significant, indicating that a response of 51/147 is unlikely under the null hypothesis. Iteratively toggling the number of responders from 51 down and performing a one-sided exact binomial test after each toggle reveals that the p-value crosses the 0.05 threshold at 29/147, where it is 0.072. The number of toggles is 22; therefore, DFI = 22. DFQ = DFI/N = 22/147 = 0.150 = fr.

The summary statistic fr = 0.15, which is considered at least moderately stable; 15% of outcomes would need to change to flip statistical significance.

#### Robustness (Proportion-NBF)

Scale factor S = √[p_0_(1 − p_0_)/n] = √[0.15 × 0.85/147] = 0.0295. Distance from neutrality = |p̂ − p_0_| = 0.1969. nb = 0.1969/(0.1969 + 0.0295) = 0.870

The summary statistic nb = 0.87 indicates that the result shows clear separation from the 15% benchmark.

#### Triplet (p, fr, nb)

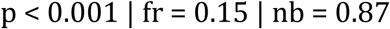

All three metrics align to support strong, reliable evidence of efficacy.

### Example 2 — Discordant pattern 1: significant but fragile evidence (two-arm binary outcome)

The OPTIMISTIC trial randomized 194 patients with acute ischemic stroke and anterior large-vessel occlusion undergoing endovascular thrombectomy to intravenous tirofiban (n=99) or control (n=95). The primary efficacy outcome was first-pass recanalization without symptomatic intracranial hemorrhage (intention-to-treat population) [17]. Outcomes are shown in Table 1.

**Table 1.** First-pass recanalization without symptomatic intracranial hemorrhage.

|  | Success | Failure | Total |
| --- | --- | --- | --- |
| Tirofiban | 64 | 35 | 99 |
| Control | 46 | 49 | 95 |
| Total | 110 | 84 | 194 |

#### Significance

Two-sided Fisher’s exact test: p = 0.0296. The results are statistically significant, supporting the alternative hypothesis that an effect exists.

#### Fragility (GFQ)

The GFQ is utilized for two-arm binary outcome studies. The shortest path-independent sequence of cell toggles that raises p above 0.05 requires two cell moves so the GFI = 2. GFQ = GFI / N = 2 / 194 = 0.010 = fr. The summary statistic fr = 0.010, indicating that the result is very fragile: only 1% of outcome changes would be required to flip the p-value from significant to nonsignificant.

#### Robustness (RQ)

The RQ determines distance from neutrality for two-arm binary outcome studies.

The sum of absolute observed-expected deviations under independence = 31.464 (average 7.866 per cell). RQ = 7.866 / (194/4) = 0.162 = nb.

The summary statistic nb = 0.16 indicates a moderate separation from independence.

#### Triplet (p, fr, nb)

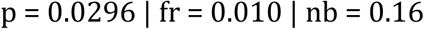

The result is formally significant but very fragile, with moderate separation from the neutrality boundary of independence. This is a classic example of statistical significance that does not withstand minimal perturbation and is only moderately separated from neutrality.

### Example 3 — Discordant pattern 2: non-significant but not neutral evidence (continuous two-group outcome)

A randomized pilot trial evaluated watermelon consumption in 39 adults with elevated blood pressure (12 in the control group, 15 in the 1 cup of watermelon per day group, and 12 in the 2 cups of watermelon per day group). The primary outcome was the change in ambulatory systolic blood pressure in individuals with elevated blood pressure [18]. For this example, we compare the systolic blood pressure of the control group with the 2 cups of watermelon per day group at week 4.

#### Significance

The Welch two-sided t-test is utilized. Control group: mean = 130.2 mm Hg, SD = 13.05. Watermelon group: mean = 124.9 mm Hg, SD = 13.05. Mean difference (control − watermelon) = 5.3 mm Hg. Observed |t| = 0.995 (df ≈ 22). Two-sided critical value at α = 0.05 = 2.074. Welch t-test (two-sided): t = 0.995, df ≈ 22, p = 0.331. The observed results are nonsignificant, supporting the null hypothesis (no effect).

#### Fragility (CFQ)

For comparing two continuous outcomes, the CFQ is utilized. The observed Welch |t| = 0.995. The two-sided critical t-value at α = 0.05, df = 22 = 2.074. The continuous fragility scale = |0.995 − 2.074| = 1.079. CFQ = 1.079 / (1 + 1.079) = 0.519 = fr

The summary statistic fr = 0.52 indicates that the non-significant p-value is stable.

#### Robustness

The Meaningful Change Index (MeCI) is the NBF metric utilized for continuous outcomes. The Weighted midpoint of the two means = 127.55 mm Hg. Distance from neutrality D = 2.65 / √(13.05² + 13.05²) = 0.1436. MeCI = 0.1436 / (1 + 0.1436) = 0.126. nb = 0.13.

The summary statistic nb = 0.13 indicates a moderate separation from neutrality (equality of means).

#### Triplet (p, fr, nb)

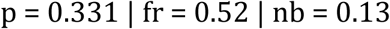

The non-significant result is highly stable (non-significance does not flip with small changes in the data). Yet, the observed means remain moderately separated from perfect equality, suggesting that an actual effect may exist. This illustrates a trial that neither proves nor disproves a meaningful effect.

A summary of the worked examples is shown in Table 2. Provisional interpretation thresholds are shown in Table 3. Note that the suggested interpretational thresholds require further empirical validation before adoption.

**Table 2.**
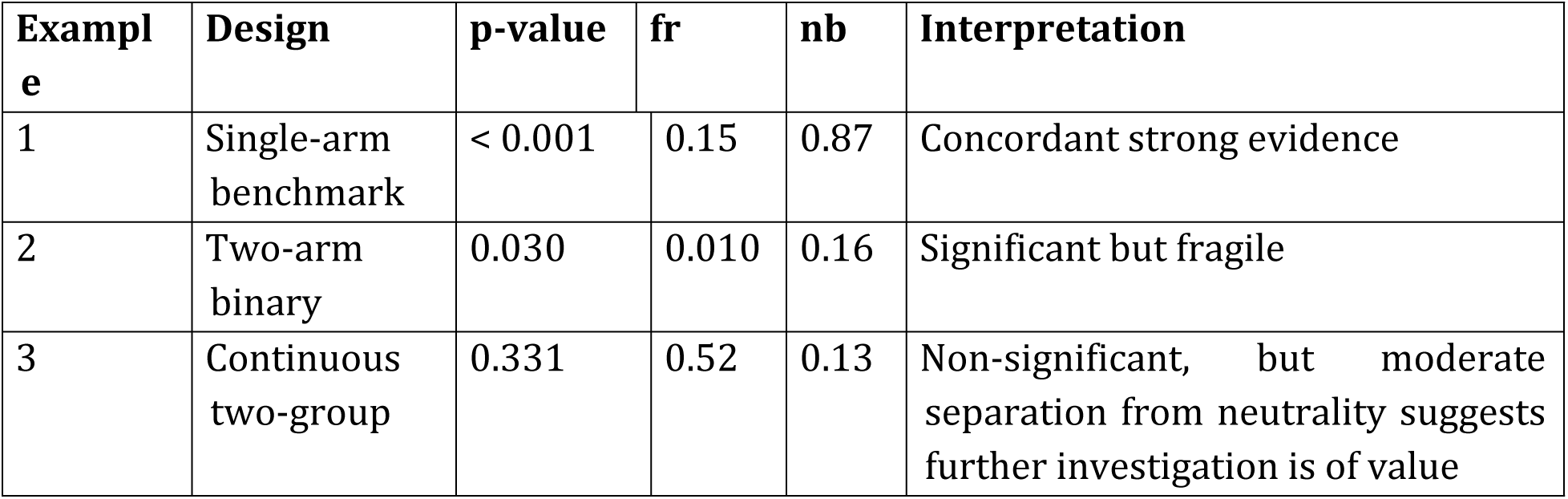
The p–fr–nb triplet across three archetypal clinical scenarios.

| Example | Design | p-value | fr | nb | Interpretation |
| --- | --- | --- | --- | --- | --- |
| 1 | Single-arm benchmark | < 0.001 | 0.15 | 0.87 | Concordant strong evidence |
| 2 | Two-arm binary | 0.030 | 0.010 | 0.16 | Significant but fragile |
| 3 | Continuous two-group | 0.331 | 0.52 | 0.13 | Non-significant, but moderate separation from neutrality suggests further investigation is of value |

**Table 3.** Provisional interpretation thresholds for fragility (fr) and robustness (nb)

| Metric | Range | Interpretation | Meaning |
| --- | --- | --- | --- |
| $fr < 0.05$ | Very fragile | Statistical conclusion flips with minimal data changes | Unstable finding; should not be trusted alone |
| $0.05 \leq fr \leq 0.10$ | Fragile | Classification is sensitive to modest changes | Requires caution; may not reproduce |
| $fr > 0.10$ | Stable | Statistical decision is unlikely to reverse under reasonable data perturbations | The p-value classification is relatively stable and resistant to small changes |
| $nb < 0.10$ | Near neutrality | Observed effect sits close to the no-effect boundary | Data consistent with no meaningful effect |
| $0.10 \leq nb \leq 0.25$ | Moderate separation | Observed effect shows measurable separation from neutrality | Data suggestive but not definitive |
| $nb > 0.25$ | Clear separation | The observed effect is far from neutrality | Strong evidence against neutrality within observed data |

Note: thresholds are provisional, context-dependent, and require empirical validation.

## Discussion

The reproducibility crisis in biomedical science has repeatedly shown that statistically significant results can be fragile or sit near the neutrality boundary [19]. In contrast, non-significant results can mask real biological separation [20]. Conventional reporting—p-value plus effect size—cannot distinguish these scenarios. The p–fr–nb triplet resolves this limitation by separating three orthogonal questions:

1. Is the result statistically significant? (p)
2. Is that significance classification stable? (fr)
3. Are the observed data geometrically far from therapeutic neutrality in units of sampling variability? (nb)

Robustness (nb) is particularly valuable when the effect size is small or complex to interpret clinically. A large nb provides strong evidence within the observed dataset that the mean or proportion lies far from neutrality—even when p is non-significant, fragility of non-significance is high, and the absolute effect size appears trivial. nb does not claim that the true population parameter is non-neutral; rather, it indicates that the *observed data* are highly inconsistent with the no-effect state.

Two theoretical examples illustrate why nb is indispensable:

### Example A

Two hypertension trials both report a statistically significant one mmHg SBP reduction (p << 0.001, fr = 0.95, and similar effect-size measures such as Cohen’s d ≈ 0.08).

- Trial 1: nb = 0.99 → The group means are 99% of the maximum possible separation given the observed variability; the data strongly contradict neutrality.
- Trial 2: nb = 0.10 → The means sit almost on the neutrality line; the one mmHg could be noise despite similar p, fr, and effect size.

Effect size and the p–fr pair treat these trials identically (“tiny, reliable effect”); nb distinguishes a meaningful shift in the observed data from an artifact.

### Example B

Two trials show a non-significant three mmHg reduction (p ≈ 0.06; f = 0.90; Cohen’s d ≈ 0.30).

- Trial 1: nb = 0.62 → the means are substantially separated from equality; the data suggest a genuine directional shift, but the study was slightly underpowered.
- Trial 2: nb = 0.07 → the means are close to neutrality; no real effect is visible in the observed data.

Again, p, fr, and effect size cannot separate these scenarios, but a large nb instantly identifies the trial that merits immediate follow-up.

The worked examples confirm these patterns in published data. In the single-arm AML trial, p = 2.65 × 10⁻⁹, fr = 0.15, and nb = 0.87 jointly support strong evidence against neutrality. In the stroke thrombectomy trial, significance is achieved (p = 0.030), but it is very fragile (fr = 0.010) and has only moderate robustness (nb = 0.16). This is a classic over-called finding. In the watermelon blood-pressure pilot, the non-significant result (p = 0.331) is stable (fr = 0.52), yet moderately separated from neutrality (nb = 0.13). This indicates that the means are not tightly clustered at neutrality, and that the trial does not convincingly prove or disprove a meaningful effect.

As biomedical research increasingly relies on surrogate endpoints, combination therapies, and meta-analyses of small effects, the ability of nb to indicate when the observed data lie meaningfully away from neutrality, despite modest effect sizes or non-significant p-values, becomes decisive. The p–fr–nb triplet—significance, stability, and geometric distance from neutrality—provides the complete, clinician-intuitive summary that p-value plus effect size alone cannot deliver. Routine reporting of all three metrics will strengthen interpretation, reduce over- and under-calling of results, and accelerate reproducible clinical science.

## Limitations

Several limitations should be acknowledged. First, all calculations rely on published summary statistics rather than individual-level data. Summary-level reporting can obscure subgroup heterogeneity, interaction effects, rounding artifacts, or variance structures that influence fragility or robustness. Second, fragility (fr) and robustness (nb) are descriptive metrics; they do not currently incorporate uncertainty intervals or claim to estimate the true population effect. Third, although nb provides a standardized 0–1 scale of distance from neutrality, correct interpretation requires specifying the appropriate neutrality boundary for the design, such as independence, equality of means, or a historical benchmark.

In addition, global fragility (GFI/GFQ) can become computationally intensive for large or complex contingency tables. In these cases, reversion to the Fragility Index and Modified-arm Fragility Quotient may be helpful [11,21]. Also, nb complements but does not replace traditional effect-size measures, which remain essential for understanding clinical magnitude. Finally, broader validation across meta-analyses and diverse clinical areas will help refine practical thresholds and establish consensus interpretation guidelines.

## Conclusion

Reporting statistical significance, fragility, and robustness together provides a multidimensional view of clinical evidence that cannot be captured by p-values or effect sizes alone. The p–fr–nb triplet separates three fundamental aspects of inference: whether an effect is statistically significant, whether that classification is stable, and how far the observed data lie from the no-effect state. This framework applies uniformly across single-arm benchmarks, binary treatment comparisons, and continuous outcomes, offering a generalizable and intuitive way to quantify evidence strength.

By aligning statistical interpretation with the realities of reproducibility and clinical decision-making, the p–fr–nb triplet enhances transparency and reduces misclassification of borderline findings. Incorporating fragility and robustness metrics alongside conventional statistics has the potential to substantially strengthen the evidentiary foundation of biomedical research and improve clinical judgment in the presence of small, uncertain, or heterogeneous effects.

## Data Availability

All relevant data are within the manuscript and its Supporting Information files. The software utilized for calculations is in the Zenodo repository: https://doi.org/10.5281/zenodo.17254763

